# Adverse respiratory events during treatment with gabapentin and opioids among older adults with spine-related conditions: a propensity-matched cohort study in the US Medicare population

**DOI:** 10.1101/2024.09.30.24314627

**Authors:** Laura S. Gold, Patrick J. Heagerty, Ryan N. Hansen, Janna L. Friedly, Richard A. Deyo, Michele Curatolo, Judith A. Turner, Sean D. Rundell, Jeffrey G. Jarvik, Pradeep Suri

## Abstract

**Background Context:** Recent work indicates no increased mortality risk with concurrent gabapentin and opioid use when using an active comparator control design. However, concurrent gabapentin and opioid prescriptions have been associated with greater risk of respiratory depression in some studies.

**Purpose:** To compare the risk of respiratory events among Medicare enrollees with spine-related diagnoses treated with gabapentin + opioids vs those treated with tricyclic antidepressants (TCA) or duloxetine + opioids. We hypothesized that enrollees treated with gabapentin + opioids would have increased risk of adverse respiratory events compared to those treated with an active control + opioids.

**Study Design/Setting:** Propensity score-matched cohort study with an incident user, active comparator (TCA/duloxetine) control design. The primary analysis included those who concurrently (within 30 days) filled ≥1 incident gabapentin + ≥1 opioid or ≥1 incident TCA/duloxetine + ≥1 opioid prescription.

**Patient Sample:** U.S. Medicare beneficiaries with spine-related diagnoses 2017-2019. People treated with gabapentin + opioids (n=66,580) were matched on demographic and clinical factors to people treated with TCAs/duloxetine + opioids (n=66,580).

**Outcome Measures:** Time to a composite respiratory outcome consisting of mechanical ventilation, intubation, respiratory failure, pneumonia, or acute respiratory distress syndrome.

**Methods:** Cox proportional hazard regression was used to estimate adjusted hazard ratios (aHRs) and 95% confidence intervals (95% CIs).

**Results:** Among 133,160 Medicare enrollees (median age 73.3 years; 66.7% female), 6089 (4.6%) experienced respiratory events before the end of follow-up. A total of 3297 (5.0%) of people who were treated with gabapentin + opioids (median initial dose/day of gabapentin was 338 mg) had respiratory events compared to 2792 (4.2%) of those treated with an active control + opioids. The increased risk in those treated with gabapentin + opioids was statistically significant after adjustment (HR 1.14; 95% CI 1.09, 1.20; p<0.0001). The most common respiratory events were pneumonia (3.5% of people in the gabapentin + opioids group versus 3.0% of people in the TCA/duloxetine + opioids group) and respiratory failure (2.2% in the gabapentin + opioids group versus 1.8% in the TCA/duloxetine + opioids group). Results were similar in analyses (a) restricted to ≤30-day follow-up and (b) that required ≥2 fills of each prescription.

**Conclusions:** While recent work has indicated no increased mortality risk with concurrent gabapentin and opioid use, the current findings suggest clinicians should exercise caution in prescribing gabapentin to people experiencing pain who are also being treated with opioids, due to the potentially increased risk of respiratory events.

## Introduction

Given recommendations from the Centers for Disease Control and Prevention [1] and the American Geriatrics Society [2] for caution in prescribing opioids for pain, prescriptions of alternative medications to treat pain, such as gabapentin, have increased [3, 4]. In a large randomized controlled trial of patients with back pain in primary care [4], an epidemiologic intervention that decreased opioid use also resulted in a higher number of prescriptions of tricyclic antidepressants (TCAs) and non-steroidal anti-inflammatory drugs (NSAIDs). Combinations of medications are also commonly used to treat chronic pain in the United States.[5, 6]

Although use of gabapentin alone is generally considered safe, some studies show associations with adverse respiratory events when it is used in combination with opioids [7]. One study found an association between gabapentin prescriptions and chronic obstructive pulmonary disease (COPD) exacerbations [8]. Other studies have found that perioperative use of gabapentinoids for pain is associated with postoperative respiratory depression [9], mechanical ventilation [10], and pneumonia compared to patients not exposed to gabapentin perioperatively [11]. On the other hand, some studies have found advantages associated with gabapentin use compared to a placebo, including reduced total morphine consumption when gabapentin was used preoperatively [12], and reduced fentanyl consumption and shorter durations of mechanical ventilation when gabapentin was used in an intensive care population [13].

While prior studies have found increased risks of respiratory events among people treated with gabapentin + opioids compared to opioids alone [14], these results may be explained by confounding due to the need for a second pain medication. That is, people taking both gabapentin and opioids may on average have more severe pain and comorbidities than those taking opioids alone. These factors alone might account for greater risk of adverse events. Such confounding may be mitigated in research by using an active control medication as a comparator. TCAs and duloxetine, - like gabapentin, are commonly prescribed to treat chronic pain. Therefore, patients receiving TCAs or duloxetine (TCA/duloxetine) are a reasonable active control group for patients receiving gabapentin.

In a recent study of the entire Medicare population with spine-related conditions which used TCAs/duloxetine as an active control group [Gold et al., in press at PAIN, [15], we found no increased risk of mortality among Medicare enrollees treated with gabapentin + opioids compared to those treated with TCAs/duloxetine + opioids. However, a secondary analysis found that people treated with gabapentin + opioids were at a slight but statistically significant increased risk of experiencing major medical complications. This raised the question of whether adverse respiratory events might explain the increased risk of major medical complications that were observed in people treated with gabapentin + opioids.

The purpose of this study was to compare time to adverse respiratory events among Medicare enrollees with spine-related conditions who were treated with gabapentin + opioids compared to those treated with TCA/duloxetine + opioids. We hypothesized that enrollees treated with gabapentin + opioids would have increased risk of adverse respiratory events compared to those treated with an active control + opioids.

## Methods

This manuscript adheres to RECORD-PE[16] and START-RWE[17] guidelines for reporting. This study was determined to be exempt from review by the University of Washington Institutional Review Board.

### Study Population

We identified enrollees from the 100% sample of Centers for Medicare and Medicaid Services (CMS) beneficiaries from 2017-2019 who had International Classification of Diseases, 10th Revision, Clinical Modification (ICD-10-CM) inpatient or outpatient diagnosis codes [18] for spine-related conditions [19-21] (e.g., neck pain, low back pain, spinal stenosis, radiculopathy, others; eTable 1). We excluded individuals with the following on or before their “index” fill date of gabapentin, TCA/duloxetine, or opioids: age <65 years; not enrolled in CMS Part A, Part B and Part D for ≥1 year before index; enrolled in a health maintenance organization (HMO) health plan (because we did not have access to data from these plans); or diagnoses of “red flag” spine conditions (e.g., fracture) or seizure disorders (eTable 2).

### Medication Exposure Groups

Medication exposures (filled prescriptions only) were ascertained from Medicare Part D claims. People with incident gabapentin fills (no gabapentin fills during the previous year) (eTable 3) and concurrent opioid fills within 30 days were compared to an active control group of people who had incident TCA or duloxetine fills (no TCA/duloxetine fills in prior year) and concurrent (within 30 days) opioid fills (eFigures 1a-1b). Selective serotonin reuptake inhibitors and serotonin-noradrenaline reuptake inhibitors (besides duloxetine) were not included in the active control group because they are less commonly prescribed for pain [22]. To ensure that opioids and the active comparator treatment were likely to have been taken concurrently, all beneficiaries included in our analysis had an “index fill” and a “qualifying fill.” For people in the gabapentin + opioids group, (1) the index fill could have been the calendar date of an opioid fill and the qualifying fill was the date on which they filled their first gabapentin prescription, or (2) the index fill could have been the calendar date on which they filled their first gabapentin prescription and the qualifying fill was the date of their opioid prescription fill. Similarly, for people in the TCA/duloxetine + opioids group, (1) the index fill could have been the calendar date on which a person filled an opioid prescription and the qualifying fill was the date on which they filled their first TCA/duloxetine prescription, or (2) the index fill could have been the calendar date on which they filled their first TCA/duloxetine prescription and the qualifying fill was the date of their opioid fill.

The primary analysis examined people who filled ≥1 gabapentin + ≥1 opioid prescription within a 30-day period compared to those who filled ≥1 TCA/duloxetine + ≥1 opioid prescription within 30 days. This analysis was designed to capture early respiratory events based on the putative mechanism of risk (that the medications taken concurrently would increase the likelihood of respiratory depression) [7]. We also conducted a secondary analysis designed to increase the likelihood that people actually consumed the medications from their filled prescriptions. The typical period of time between refills for gabapentin is 1-3 months [23], so this secondary analysis examined people with ≥2 gabapentin fills + ≥2 opioid fills within a 120-day exposure period and compared them to those with ≥2 TCA/duloxetine fills + ≥2 opioid fills in 120 days (eFigures 1c-1d).

We also calculated the mean daily doses of opioids (in milligram morphine equivalents [MMEs][24]), gabapentin, duloxetine, and each TCA (in milligrams (mg)) for the index fills as well as through the end of follow-up. For the dosages from index through the end of follow-up, we compared the dosages in each drug category among those who did and did not have respiratory events.

### Outcomes and Censoring Variables

Beginning from the day of the qualifying fill, we assessed occurrence of any of five respiratory events at any time and at 30 days, at 1 year, and at 2 years after the qualifying date: intubation, mechanical ventilation, respiratory failure, pneumonia, or acute respiratory distress syndrome (ARDS) (eTable 4). Because the posited mechanism of action of gabapentin + opioids is that the medications may increase risk of respiratory depression, [14] we chose respiratory events associated with respiratory depression and its sequelae as the main study outcomes. We did not include respiratory events that are not associated with respiratory depression (e.g., pulmonary embolism).

Censoring events were fills of benzodiazepine or pregabalin after the qualifying fill (because concurrent use of benzodiazepines independently increases respiratory depression among opioid users [25] and pregabalin is pharmacologically similar to gabapentin [26]); no opioid refill for >45 days; TCA/duloxetine fill in comparison groups treated with gabapentin; no gabapentin refill for >180 days in comparison groups treated with gabapentin; gabapentin fill in comparison groups treated with TCA/duloxetine; no TCA/duloxetine refill for >180 days in comparison groups treated with TCA/duloxetine; or the end of the study (December 31, 2019), whichever occurred first.

### Statistical Analysis

To increase internal validity and limit confounding, we used two approaches to ensure that the TCA/duloxetine group was as comparable as possible to the gabapentin group in terms of covariates that could have caused residual confounding. First, we included a wide range of variables in the logistic regression used for propensity score 1:1 matching based on clinical knowledge and prior literature (eTable 5). In particular, given that duloxetine and TCAs are common treatments for mental health conditions, and the same is true for gabapentin (although to a lesser extent), we included several variables indicative of inpatient or outpatient psychiatric diagnoses in the propensity score, including whether people had diagnoses of alcohol abuse and opioid use disorder in the year prior to index, as well as the counts of the number of visit dates (as surrogates for severity) with diagnoses of mood disorders, generalized anxiety disorder (GAD), and post-traumatic stress disorder (PTSD) in the year prior.

Second, because we expected certain variables that occurred between the index and qualifying fills to be strongly related to whether people received the prescriptions of interest *and* to respiratory events, we matched on them “exactly” in addition to matching on the propensity score. These included (1) average daily MME [24] of opioid prescriptions filled on/between the index and qualifying fill dates, matched within 9 categories (>0 to 2.1; >2.1 to 5.3; >5.3 to 10.0; >10.0 to 15; >15 to 20; >20 to 30; >30 to 40; >40 to 64, and >64) based on MME distribution; (2) the length of time between the index and qualifying fills to make the timing of concurrent opioid treatment comparable between groups; (3) the presence of a respiratory event between the index and qualifying fills (eTable 4); and (4) the presence of non-respiratory complications between index and qualifying fills (eTable 6).

We calculated descriptive statistics (numbers, proportions, medians, interquartile ranges (IQRs), and standardized mean differences) to ensure the matched groups were similar. Kaplan-Meier curves were plotted to graphically assess the proportional hazards assumption, which was met for all analyses. Cox proportional hazard regression was used to estimate adjusted hazard ratios (aHRs) and 95% confidence intervals (95% CIs). All regression models were adjusted for the variables in the propensity score model (eTable 5). Two-sided p-values of <0.05 were considered statistically significant and analyses were performed using SAS Enterprise Guide version 7.15 and SAS for Windows version 9.4, SAS Institute Inc., Cary, NC.

## Results

The flow of Medicare beneficiaries in the primary analysis is shown in Figure 1. Of the initial 525,884 people who were eligible for matching, 66,580 people treated with gabapentin + opioids matched to 66,580 people treated with TCA/duloxetine + opioids. An abbreviated list of cohort characteristics is shown in Table 1 and a complete list (with standardized mean differences) is shown in eTable 7. The median (range) follow-up time was 45 (1-1093) days. Although the unmatched treatment groups differed with respect to various characteristics (eTable 7), the matched groups were closely comparable (Table 1 and eTable 7). All subsequent results refer to the matched groups. The majority of people were female, white, and had a mean (standard deviation) Charlson comorbidity score of 2.7 (2.7). In both treatment groups, mental health diagnoses and fills of medications besides opioids, gabapentin, TCA, or duloxetine that are often prescribed for pain were common. In the TCA/duloxetine group, 62% of Medicare beneficiaries filled duloxetine for their index fill and 38% filled a TCA. The median (IQR) doses of the index fills were: 300 mg (300, 900) for gabapentin, 30 mg (30, 60) for duloxetine and 25 mg (10, 30) for amitriptyline, the most common TCA that was filled.

**Table 1.**
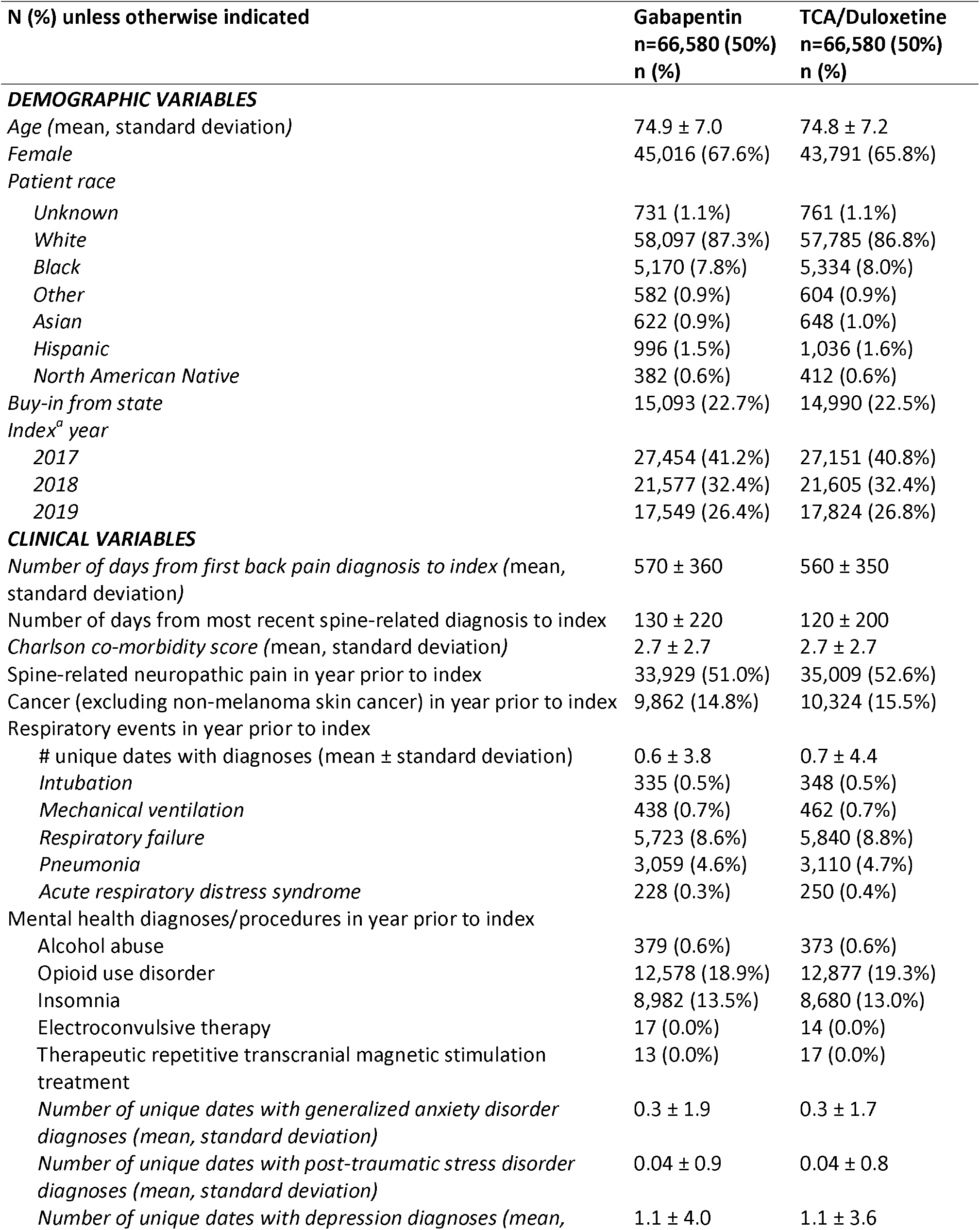
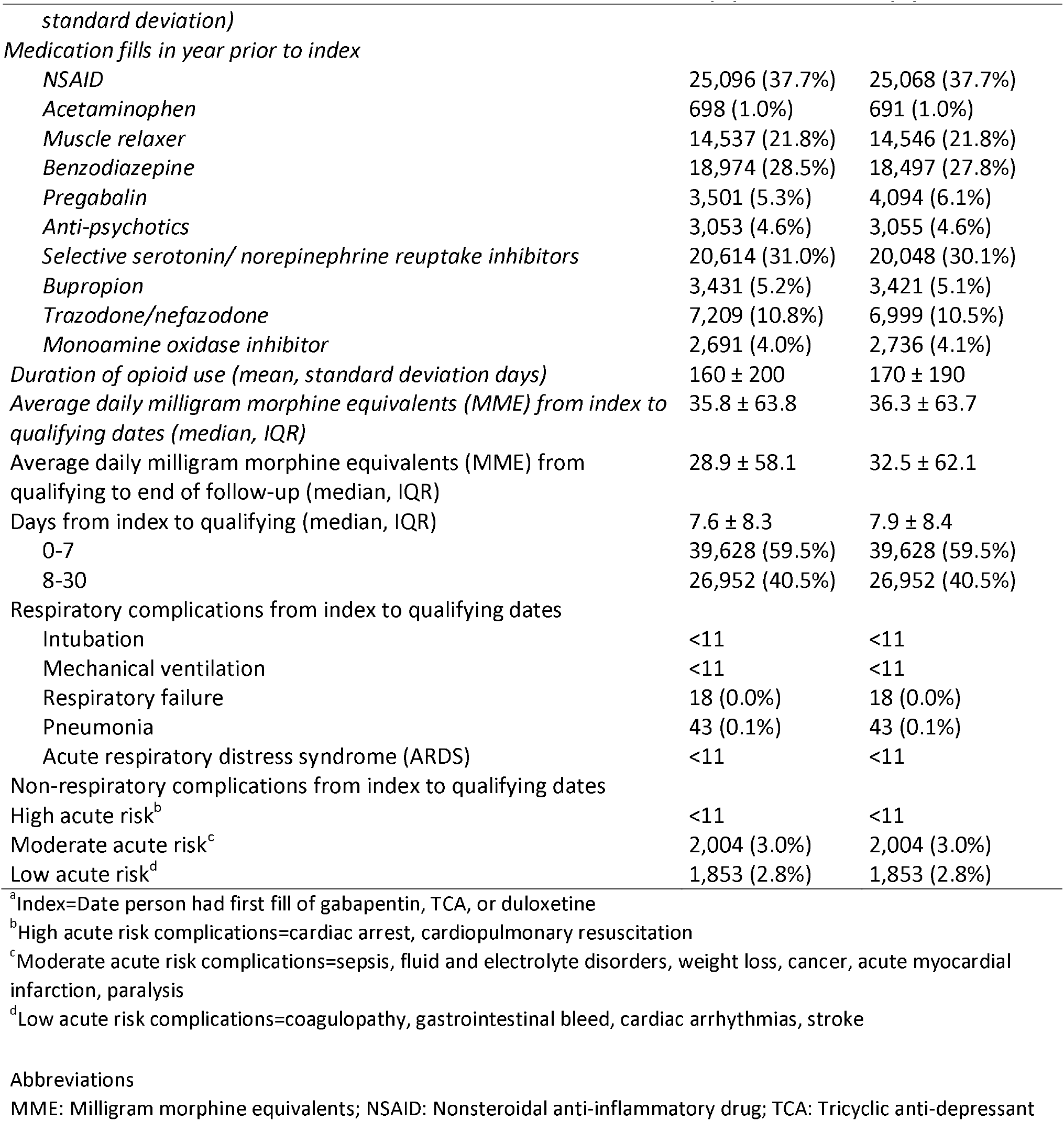
Comparison of patients who received gabapentin to those who received TCAs/duloxetine using 1:1 matching in the primary analysis (selected variables).

**Figure 1.**
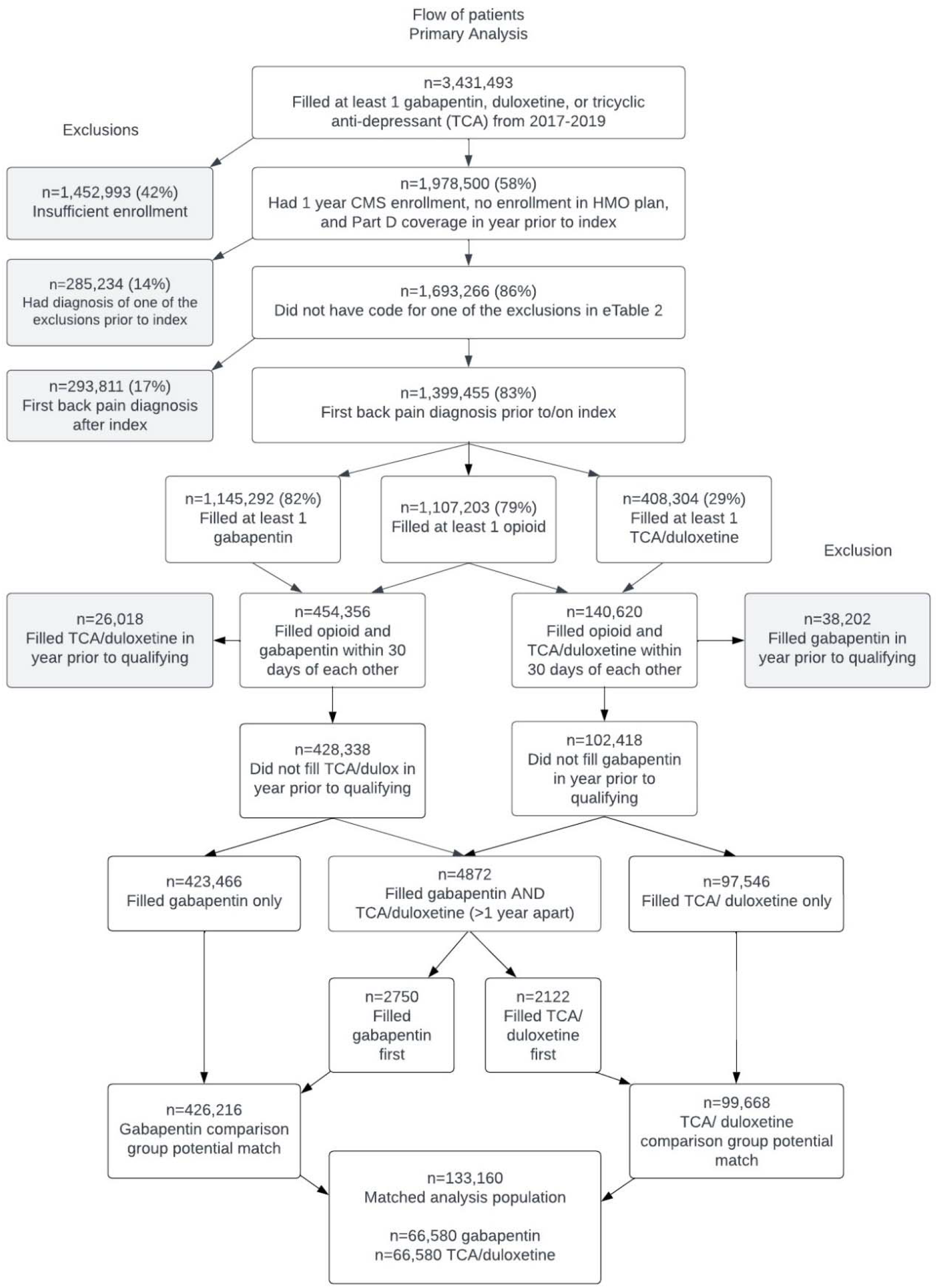
Flow of patients for the primary analysis.

We observed a statistically significant increased risk of respiratory events comparing drug treatment groups over the entire follow-up time (eFigure 2; Figure 2) (aHR, 1.14; 95% CI, 1.09, 1.20). Comparing those treated with gabapentin + opioids versus those treated with TCA/duloxetine + opioids, the proportion at risk for respiratory events was very similar at 1-month, but the difference widened by 1-year and 2-years after index (Figure 2).

**Figure 2.**
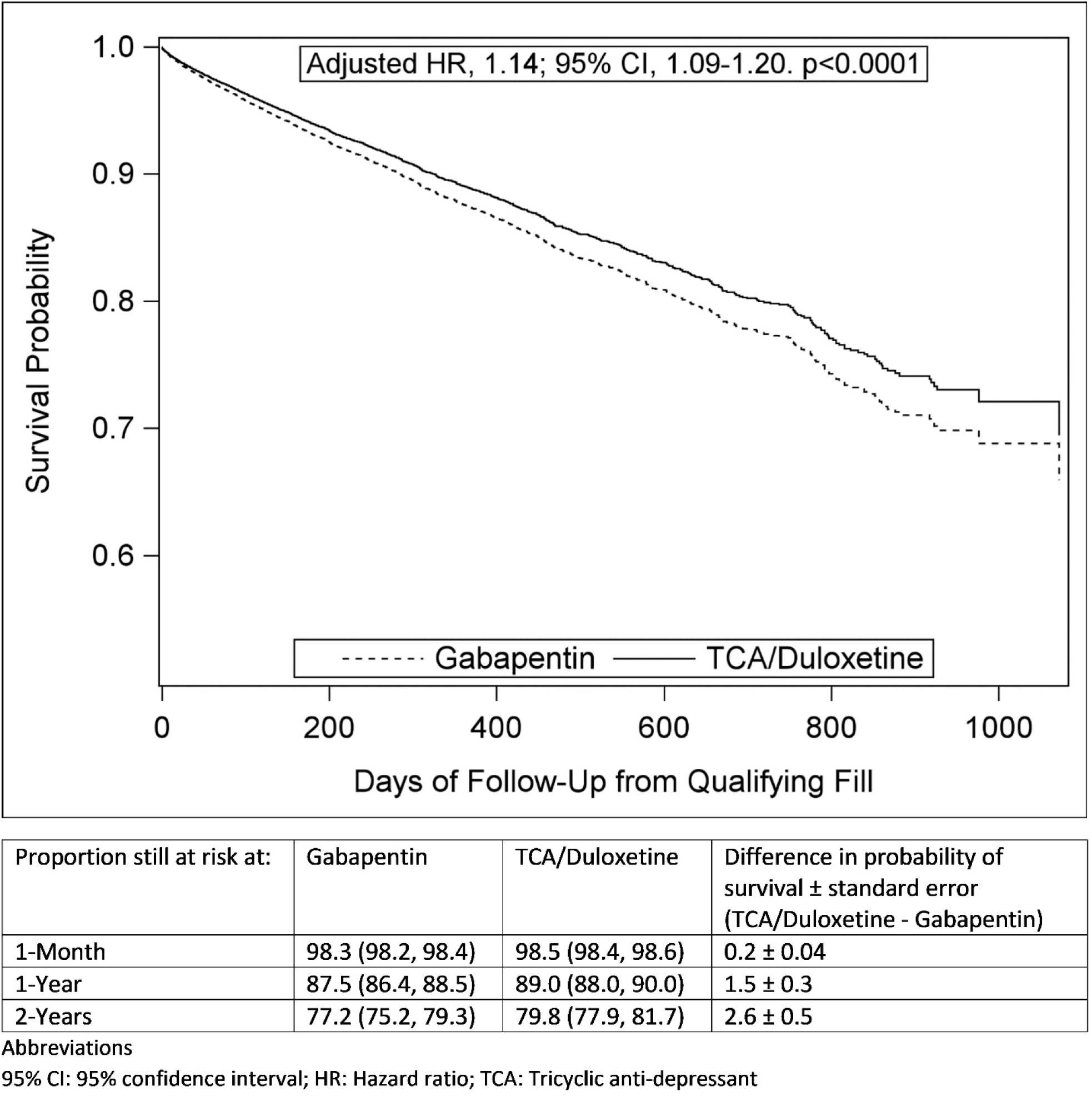
Primary analysis Cox proportional hazard curves for any-time respiratory events.

The numbers and percentages of people who experienced each respiratory event are shown in Table 2. Although respiratory events were not uncommon in both drug treatment groups, a greater proportion of people in the gabapentin group experienced a respiratory event (5.0% versus 4.2%). The most common types of respiratory events were pneumonia (3.5% of people in the gabapentin + opioids group versus 3.0% in the TCA/duloxetine + opioids group) and respiratory failure (2.2% in the gabapentin + opioids group versus 1.8% in the TCA/duloxetine + opioids group). People in the gabapentin group were somewhat more likely to have been censored because they did not refill an opioid prescription for >45 days (53.0% versus 47.3%) and less likely than people in the TCA/duloxetine + opioids group to have been censored for filling pregabalin prescriptions (2.6% in the gabapentin + opioids group versus 4.6% in the TCA/duloxetine + opioids group) (Table 2). Results were similar when we examined 30-day respiratory events (eTable 8; eFigures 3-4).

**Table 2.** Primary analysis respiratory outcomes at any time.

|  | **Gabapentin**<br>**n=66,580 (50%)**<br>**n (%)** | **TCA/Duloxetine 3**<br>**n=66,580 (50%)**<br>**n (%) 4** |
| --- | --- | --- |
| Any respiratory complication | 3,297 (5.0%) | 2,792 (4.2%) |
| Days to first respiratory complication (median, IQR) | 37 (13, 118) | 39 (12, 113) |
| Intubation | 190 (0.3%) | 175 (0.3%) |
| Mechanical ventilation | 231 (0.3%) | 214 (0.3%) |
| Respiratory failure | 1,476 (2.2%) | 1,202 (1.8%) |
| Pneumonia | 2,354 (3.5%) | 1,979 (3.0%) |
| Acute respiratory distress syndrome | 51 (0.1%) | 38 (0.1%) |
| Reasons for follow-up end |  |  |
| Disenrolled | 8,806 (13.2%) | 9,058 (13.6%) |
| No opioid refill >45 days | 35,268 (53.0%) | 31,488 (47.3%) |
| Pregabalin fill | 1,745 (2.6%) | 3,043 (4.6%) |
| Benzodiazepine fill | 11,317 (17.0%) | 11,576 (17.4%) |
| TCA/duloxetine fill in gabapentin group | 1,820 (2.7%) | -- |
| Gabapentin fill in TCA/duloxetine group | -- | 3,742 (5.6%) |
| No gabapentin refill >180 days | 4,327 (6.5%) | - |
| No TCA/duloxetine refill >180 days | -- | 4,881 (7.3%) |
| Respiratory complication | 3,297 (5.0%) | 2,792 (4.2%) |

People with respiratory events had higher median opioid doses per day in both drug treatment groups (Table 3). Among those treated with gabapentin + opioids, the median (IQR) opioid dose was 21.4 (9.2, 48.9) MME in those with respiratory events vs 11.6 (4.4, 30.0) MME in those without. Among those treated with TCA/duloxetine + opioids, the median (IQR) MME was 22.4 (9.3, 50.9) in those with respiratory events versus 13.9 (5.0, 34.0) in those without. The median dose of gabapentin in the last fill prior to the end of follow-up was higher than the median dose from index to the end of follow-up among everyone, but the difference was more pronounced among people with respiratory events compared to those without (median (IQR) 600 (300, 900) mg in those with respiratory events versus 400 (300, 900) mg in those without). Those who did not have respiratory events tended to have somewhat higher dosages of duloxetine and of each of the TCAs (Table 3).

**Table 3.** Median daily doses of opioids, gabapentin, TCA, and duloxetine, stratified by whether person had respiratory complication.

| Medication | Timing | Gabapentin |  |  |  | TCA/Duloxetine |  |  |  |
| --- | --- | --- | --- | --- | --- | --- | --- | --- | --- |
|  |  | No respiratory complication |  | Respiratory complication |  | No respiratory complication |  | Respiratory complication |  |
|  |  | n | Median (IQR) | n | Median (IQR) | n | Median (IQR) | n | Median (IQR) |
| Opioid (MME/day) | Qualifying date to EOF | 63,283 | 11.6 (4.4, 30.0) | 3297 | 21.4 (9.2, 48.9) | 63,788 | 13.9 (5.0, 34.0) | 2792 | 22.4 (9.3, 50.9) |
| Gabapentin (mg/day) | Index date to EOF | 63,283 | 340 (200, 720) | 3297 | 320 (170, 640) | -- | -- | -- | -- |
| Gabapentin (mg/day) | Last dose before EOF | 63,283 | 400 (300, 900) | 3297 | 600 (300, 900) | -- | -- | -- | -- |
| Duloxetine (mg/day) | Index date to EOF | -- | -- | -- | -- | 39,758 | 33.3 (22.3, 60.0) | 1836 | 31.0 (20.0, 50.5) |
| Amitriptyline (mg/day) | Index date to EOF | -- | -- | -- | -- | 11,798 | 22.5 (10.6, 35.7) | 463 | 19.0 (8.9, 36.4) |
| Imipramine (mg/day) | Index date to EOF | -- | -- | -- | -- | 670 | 28.9 (16.7, 53.8) | 27 | 19.2 (8.1, 49.9) |
| Nortriptyline (mg/day) | Index date to EOF | -- | -- | -- | -- | 5309 | 19.4 (10.3, 31.2) | 193 | 15.8 (9.1, 25.9) |
| Doxepin (mg/day) | Index date to EOF | -- | -- | -- | -- | 6078 | 13.3 (6.7, 30.0) | 267 | 10.6 (5.6, 26.1) |
| Other TCA<sup>a</sup> (mg/day) | Index date to EOF | -- | -- | -- | -- | 312 | 26.6 (11.8, 60.1) | 13 | 13.5 (5.1, 45.5) |

<sup>a</sup>Amoxapine, clomipramine, desipramine, maprotiline, protriptyline, trimipramine maleate
EOF: End of follow-up; IQR: Interquartile range; Mg: Milligram; MME: Milligram morphine equivalents; TCA: Tricyclic anti-depressant

Results of the secondary analysis requiring ≥2 fills of each medication of interest were similar to those of the primary analysis. A total of 165,585 people who filled ≥2 gabapentin and opioid prescriptions were eligible to be matched with 42,344 people who filled ≥2 TCA/duloxetine and opioid prescriptions; among these, 15,522 were in each matched group (eFigure 5). The groups were well-matched (eTable 9). We observed a greater proportion of respiratory events among people treated with gabapentin at any time (eTable 10) and an increased adjusted risk of a respiratory event among people treated with gabapentin + opioids (eFigures 6-7; aHR, 1.19; 95% CI, 1.08, 1.32). Results for 30-day respiratory events in the secondary analysis were similar to the primary analysis (eTable 11; eFigures 8-9).

## Discussion

In this analysis of Medicare beneficiaries with spine-related diagnoses, we found that people treated with gabapentin + opioids were at a small but statistically significant increased risk of respiratory events compared to people treated with TCA/duloxetine + opioids, with a 14% greater relative risk of a respiratory event within 1 year and a 1.5% greater absolute risk within 1 year. This relationship was consistent when we examined respiratory events within 30 days, as well as when we required people in the matched cohorts to have filled ≥2 prescriptions for gabapentin/active control + opioids and therefore were more likely to have consumed their medications.

The combination of gabapentin + opioids may lead to increased risk of respiratory events by causing respiratory depression [27-30]. This hypothesis is supported by studies comparing people exposed to gabapentin versus non-users with respect to outcomes including COPD exacerbations [9], post-operative mechanical ventilation [10], pneumonia [11], and respiratory depression [8, 29]. However, by comparing gabapentin users to non-users without an active control group [9-11], these analyses were subject to substantial residual confounding and it is possible that some unmeasured confounder rather than gabapentin was causing the respiratory events. One of these studies [8] did perform a secondary analysis with an active control group. These authors examined the risk of COPD exacerbations in people in Quebec, Canada with neuropathic or chronic pain who were treated with gabapentinoids versus NSAIDs. They found that those treated with gabapentinoids were at increased risk of COPD exacerbations. Therefore, our study is consistent with and adds to the limited previous work demonstrating increased risk of adverse respiratory events with combined use of gabapentin and opioid medication.

The current study also builds on past work by showing that the increased risk of respiratory events with combined opioid and gabapentin applies not only to select groups of people with certain medical conditions (e.g., breast cancer survivors [30], surgical patients [9], those with COPD [8]) but more broadly to people with spinal pain, the most common pain condition associated with office visits that result in opioid prescriptions [31]. This is important given that spine-related diagnoses are the most common diagnoses and are a substantial driver of increasing health care costs in the United States [32].

It is important to note that the significantly increased risk of respiratory events with concurrent gabapentin and opioid use in the current analysis was present even though the doses of gabapentin used were quite low, with a median daily gabapentin dose of about 300 mg at the time of gabapentin initiation, increasing to a 400 mg daily gabapentin dose throughout follow-up among people who did not have subsequent respiratory events and to a 600 mg daily gabapentin dose among people who *did* have subsequent respiratory events. Opioid doses from the qualifying date to the end of follow-up were also higher among gabapentin users who had subsequent respiratory events versus those who did not, which is not surprising since higher doses of opioids are associated with increased risk of respiratory depression [33]. This highlights that, among older adults who are using opioids, even low doses of gabapentin may increase the likelihood of respiratory events and supports the current recommendation from the Beers criteria [2] that the combination of gabapentin and opioids should be used cautiously in older adults, or avoided.

The current findings are important to consider in the context of our recent work using nearly identical methods that found no significant difference in the risk of mortality (OR= 0.98 [0.90, 1.06]; p=0.63) among those treated with gabapentin + opioids versus those treated with TCA/duloxetine + opioids [Gold et al. in press at *PAIN*], with a point estimate that did not suggest even a slight tendency towards a detrimental effect on mortality. Together, these results suggest the possibility that adverse respiratory events after gabapentin initiation among older adults using opioids may lead to clinical changes, such as not refilling gabapentin and opioid prescriptions in people who have had respiratory events, that prevent subsequent mortality. This underscores the importance of educating patients and caregivers about the increased risk of respiratory events with opioid use, and potential augmentation of this increased risk when initiating gabapentin, so that patients and their caregivers can be watchful for changes in respiratory status. This also underscores the importance of healthcare professionals asking about shifts in respiratory status when seeing older patients who use opioids who are returning to clinic after recently initiating gabapentin.

We emphasize strongly that the current study’s findings apply only to gabapentin treatment in the context of concurrent opioid treatment among older adults and do *not* apply to gabapentin when opioids are not being used concurrently. Future studies are needed in samples of people not taking opioids to compare rates of respiratory events and other important outcomes such as falls in those taking gabapentin versus those taking an active control medication. Also, studies conducted in broad populations who do not have pre-existing respiratory diagnoses or other major comorbidities would be invaluable.

By using an active comparator group and extensive matching in this large, representative group of older US adults, we evaluated respiratory events in cohorts that were as alike as possible except with respect to their drug treatment regimens. The active comparator group also illustrated the high frequency of respiratory events among older adults using opioids in general, even those who did not use gabapentin. Despite use of an active control group, however, it remains possible that residual confounding by factors not available in claims data such as pain severity and functional limitations remained. Because our study population included only older adults with spine-related conditions, we cannot be certain that our results would apply to younger populations or people with other pain conditions.

In summary, among older adults using opioids for spinal conditions, gabapentin initiation, even at low doses, conferred a small but statistically significant risk of adverse respiratory events compared to an active control group. Thus, healthcare professionals should carefully consider whether the benefits of initiating gabapentin in older adults who have already been prescribed opioids outweigh the risks. If gabapentin is prescribed in older adults taking opioids, our findings suggest the importance of informing patients and caregivers of the risk of respiratory events and carefully monitoring them for respiratory depression and other respiratory problems.

## Data Availability

All data produced in the present study are available upon reasonable request to the authors.

